# Association of Renin Angiotensin System Blockers with Outcomes in Patients with Covid-19: A Systematic Review and Meta-analysis

**DOI:** 10.1101/2020.05.23.20111401

**Authors:** Aakash Garg, Amit Rout, Abhishek Sharma, Brittany Fiorello, John B. Kostis

**Author notes:** **Corresponding author:** Aakash Garg, MD, Division of Cardiology, Newark Beth Israel Medical Center, (USA), @Dr_AGarg_MD. **Funding:** None.

## Abstract

**Background:** Patients with cardiovascular disease are at increased risk of critical illness and mortality from Covid-19 disease. Conflicting findings have raised concerns regarding the association of angiotensin converting enzyme inhibitors (ACEI) and angiotensin receptor blockers (ARBs) use with likelihood or severity of infection during this pandemic.

**Objective:** To study the cumulative evidence for association of ACEI/ARB use with outcomes among patients with confirmed Covid-19.

**Methods:** The MEDLINE and EMBASE databases were thoroughly searched from November 01, 2019 to May 15, 2020 for studies reporting on outcomes based on ACEI/ARB use in patients with confirmed Covid-19. Preferred reporting items for systematic review and meta-analysis guidelines were used for the present study. Relevant data was collected and pooled odds ratios (OR) with 95% confidence intervals (CI) were calculated using random-effects model.

**Main Outcome measures:** In-hospital mortality was the primary end of interest. Second end-point was severe or critical illness defined as either need for intensive care unit, invasive mechanical ventilation, or mortality.

**Results:** Fifteen studies with total of 23,822 patients (N ACEI/ARB=6,650) were included in the present analysis. Overall, prevalence of ACEI/ARB use ranged from 7.7% to 46.2% across studies. Among 10 studies, patients using ACEI/ARB had similar odds of mortality [OR 1.03 (0.69-1.55)] and severe or critical illness [1.18 (0.91-1.54)] compared to those not on ACEI/ARB. In an analysis restricted to patients with hypertension, ACEI/ARB use was associated with significantly lower mortality [0.64 (0.45-0.89)], while the odds of severe/critical illness [0.76(0.52-1.12); p=0.16] remained non-significant compared with non-ACEI/ARB users.

**Conclusion:** There is no evidence for increased risk of severe illness or mortality in patients using ACEI/ARB compared with non-users. In patients with hypertension, ACE/ARB use might be associated with reduced mortality, however these findings need to be confirmed in prospective randomized controlled trials.

## Introduction

Covid-19 pandemic affecting more than 4.5 million people across the globe has caused significant morbidity and mortality [1]. Angiotensin converting enzyme 2 (ACE-2) has been implicated in the entry of SARS-Cov-2 virus into host cells [2]. Since renin angiotensin system (RAS) antagonists have been suggested to upregulate ACE2 in few animal models, concerns have been raised that these drugs might be associated with increased risk of infection or severe disease from Covid-19 [3,4].

Patients with Covid-19 infection have a substantial prevalence of cardiovascular (CV) diseases including hypertension (HTN) and heart failure [5]. Whether such patients on ACE inhibitors (ACEIs) or angiotensin receptor blockers (ARBs) should continue these drugs has become a matter of discussion. Several studies investigating the relationship between RAS inhibitors and outcomes in patients with Covid-19 have reported conflicting results [6,7]. Accordingly, we sought to study the cumulative evidence for association of ACEI/ARB use with risk of mortality and severe illness with Covid-19.

## Methods

A comprehensive search in electronic databases (MEDLINE and EMBASE) was performed of studies published between November 01, 2019 and May 15, 2020, and reporting outcomes among Covid-19 patients on RAS inhibitors. The following key words were used for search in different combinations: “Coronavirus 2019”, “Covid-19”, “SARS-Cov-2”, “Renin angiotensin system”, “Angiotensin converting enzyme”, “Angiotensin converting enzyme inhibitors”, “ACEI”, “Angiotensin receptor blockers”; “ARB”, and “Outcomes”. Inclusion criteria were studies published in peer- reviewed journals and describing outcomes based on ACEI/ARB use.

Two reviewers (A.G. and A.R.) screened the study titles and abstracts for relevance. This was followed by full manuscript evaluation including supplementary data, if available. From individual studies, we collected baseline characteristics of Covid-19 patients including proportion of patients with HTN and those taking ACEI/ARB. The primary outcome was in-hospital mortality. Secondary outcome was severe/critical illness [need for intensive care unit, invasive mechanical ventilation, or mortality] as defined per individual study protocol.

This meta-analysis was conducted in accordance with the PRISMA guidelines [8]. We used the Cochrane review manager 5.3 for statistical analysis. Random-effects model with Mantel-Haenszel method was used to calculate the pooled odds ratios (OR) with 95% confidence intervals (CI) for each end-point. A p-value <0.05 was considered significant. Heterogeneity was examined using the I^2^ statistic. Subgroup analyses were performed to study: 1) association of ACEI/ARB use with outcomes specifically in patients with HTN; 2) independent relationship of ACEI and ARB drugs with outcomes.

## Results

As reported in Online Figure 1, initial search resulted in 306 studies that were screened at abstract level. After full text review, 15 studies were identified to report outcomes based on ACEI/ARB use in patients with confirmed Covid-19 [5-7, 9-20]. A total of 23,822 COVID-19 positive patients [n = 6,650 on ACEI/ARB, n = 17,172 not on ACEI/ARB] were included.

**Figure 1.**
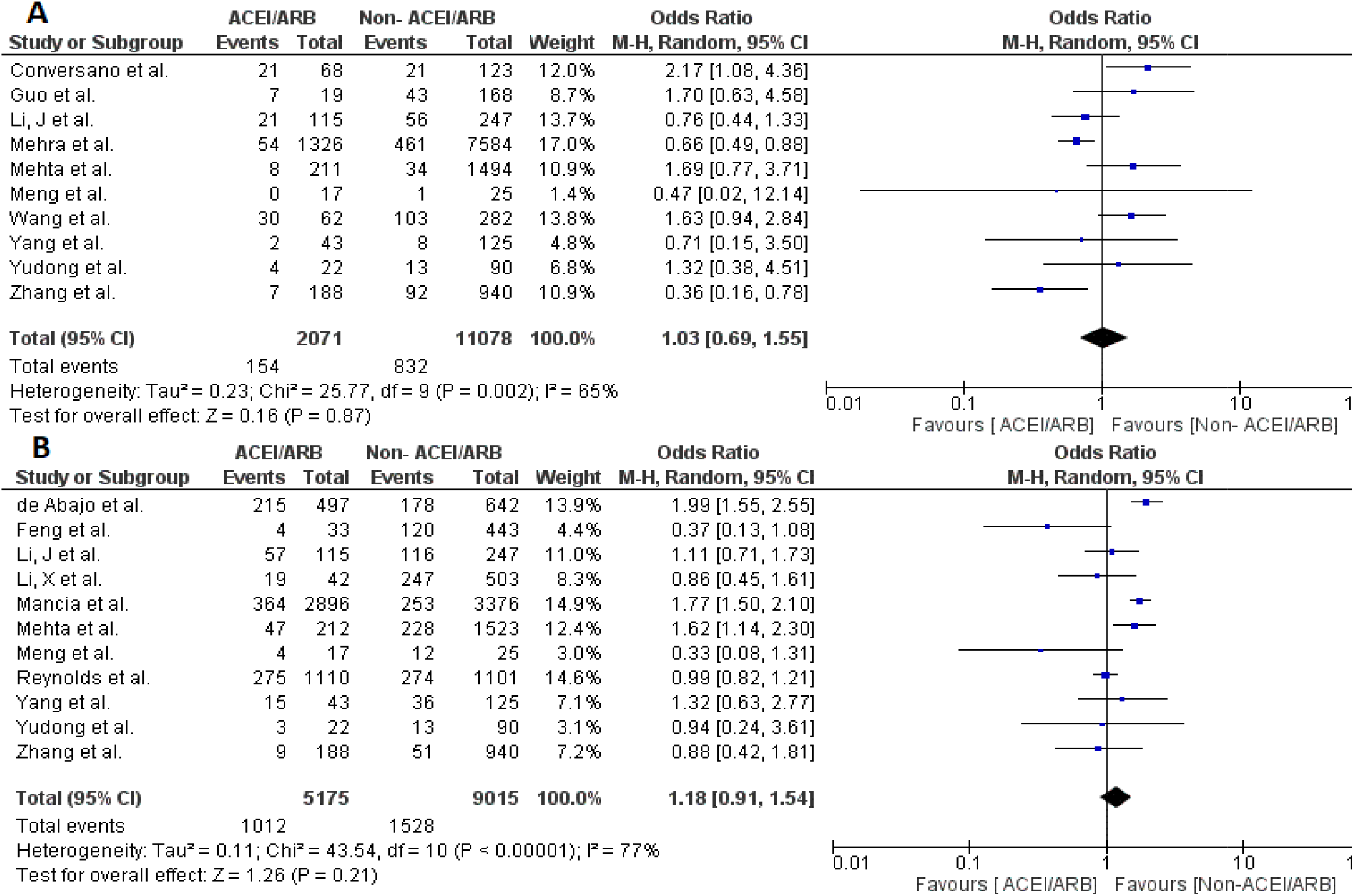
Forest plots comparing outcomes between ACEI/ARB users vs. non-ACEI/ARB users. A. Mortality. B. Severe or critical illness. ACEI: Angiotensin converting enzyme inhibitors; ARB: Angiotensin receptor blocker

Baseline characteristics and subject demographics within individual studies are described in Table 1. HTN prevalence was variable across studies (range 23.7%-100%). ACEI/ARB use ranged between 7.7% and 46.2%. Out of 15 studies, 7 studies compared outcomes with all-comers in control group [5,9-12,17,19] whereas 3 studied patients with HTN specifically [6,13,18]. Five studies reported data separately for both all-comers and HTN patients as control groups [7,14-16,20]. Outcomes based on the type of RAS inhibitors (ACEI or ARB) were reported in 9 studies [7,10-15,19-20].

**Table 1.**
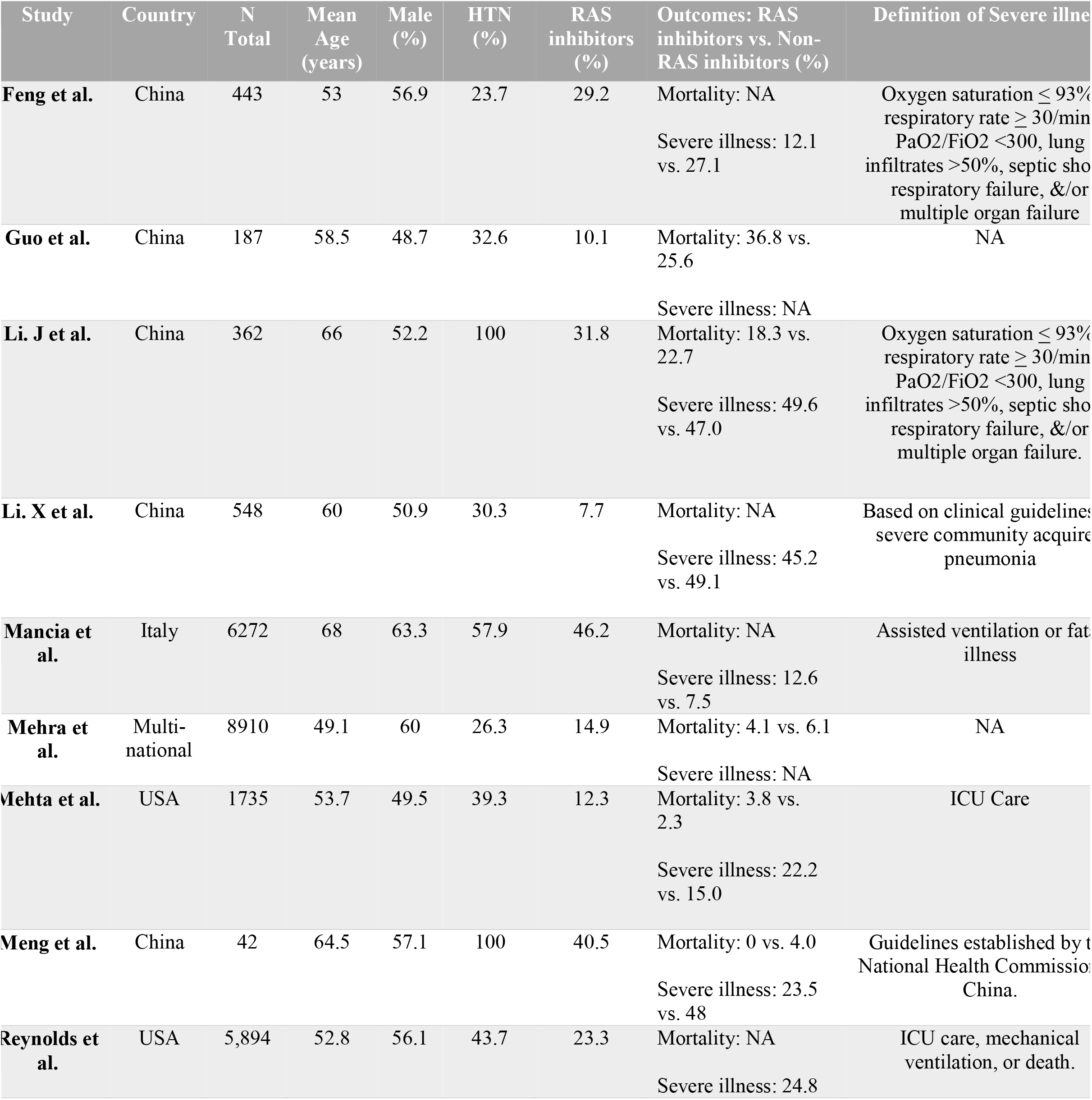
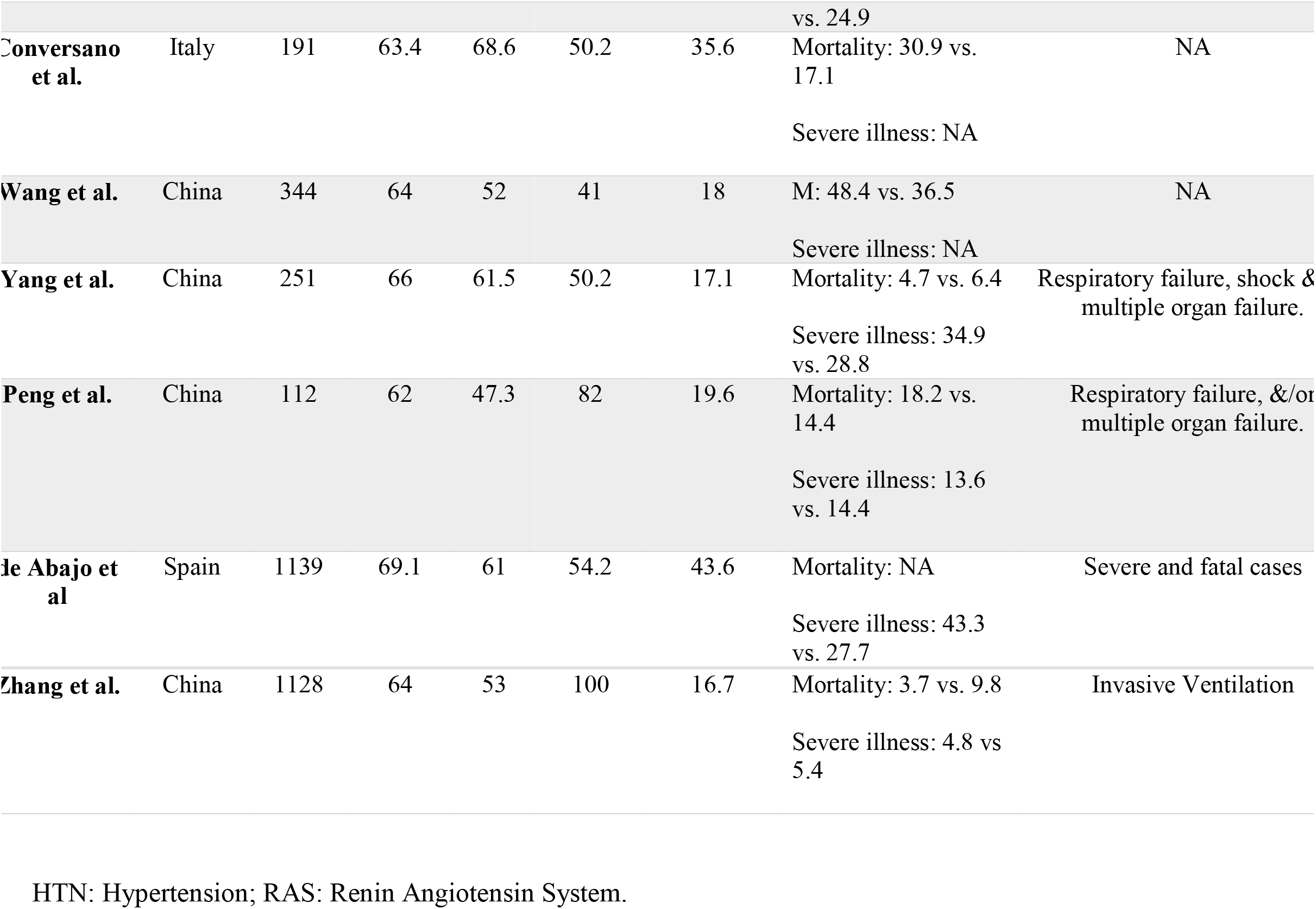
Baseline characteristics and demographics of patients in individual studies.

Comparisons of mortality and severe illness between ACEI/ARB users vs. non-users were reported in 10 and 11 studies, respectively. Compared to patients not on RAS inhibitors, patients using RAS inhibitors had similar risks for mortality [6 studies; OR 1.03 (0.69-1.55); p=0.87] and severe illness [1.18 (0.91-1.54); p=0.21] (Figure 1). Significant heterogeneity was noted across the studies (I^2^=65% for mortality and I^2^=77% for severe illness). No single study influenced results or heterogeneity in sensitivity analyses.

Subgroup analysis comprising only patients with HTN showed reduced risk of mortality in patients on ACEI/ARB when compared with those not on ACEI/ARB [0.64 (0.45-0.89); p=0.01]. In terms of severe illness, no significance was observed in trend between ACEI/ARB users vs. non-ACEI/ARB users [0.76 (0.52-1.12); p=0.16] (Figure 2).

**Figure 2.**
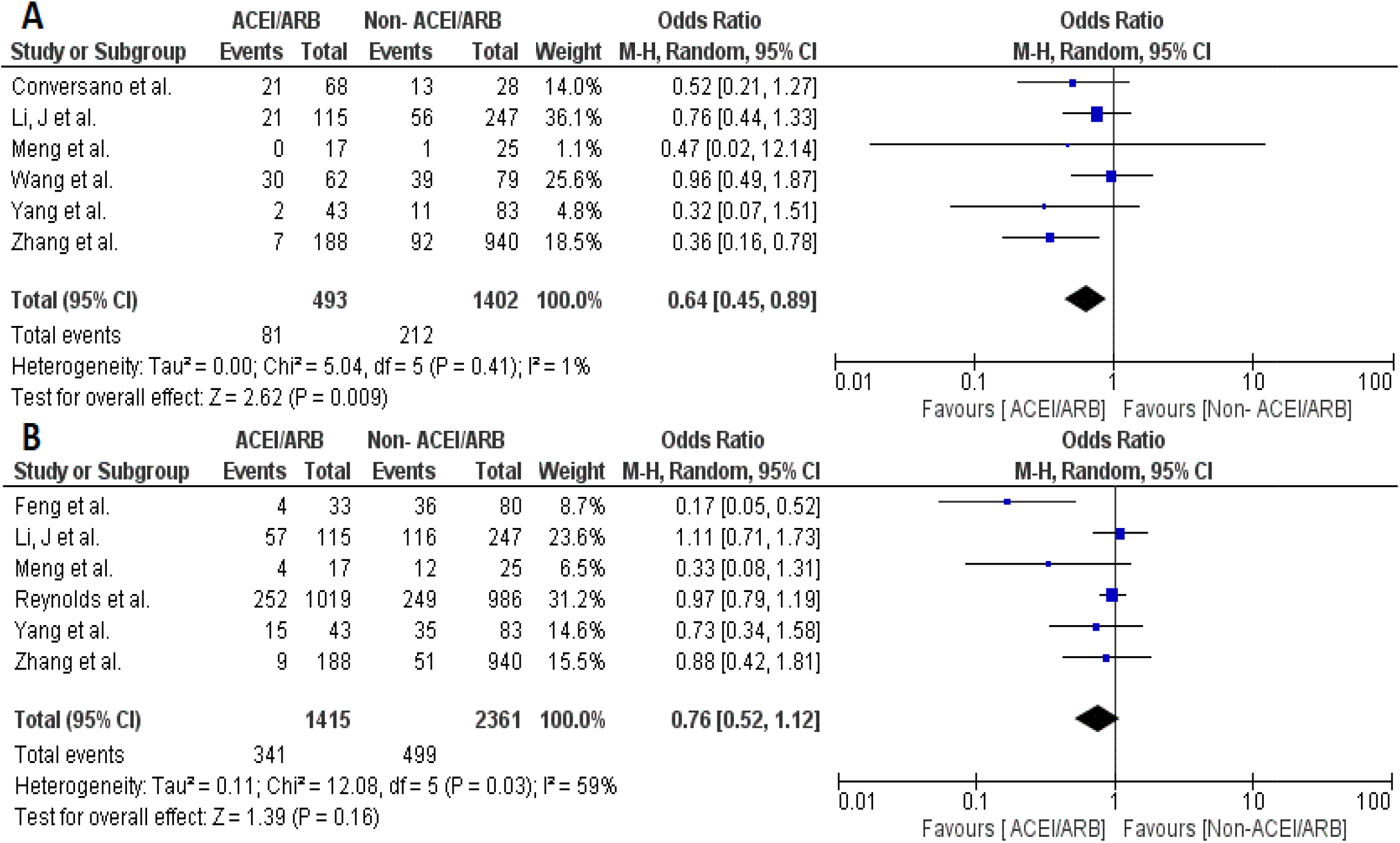
Forest plots comparing outcomes between ACEI/ARB users vs. non-ACEI/ARB users in patients with hypertension. A. Mortality. B. Severe or critical illness.

Finally, there were no differences in outcomes in subgroup analyses based on type of RAS inhibitors (ACEI vs. non-ACEI and ARBs vs. non-ARBs) (Online Figures 2-3).

## Discussion

This is the first meta-analysis on association of RAS inhibitors with outcomes in patients with Covid-19. Our main findings are that among all comers, those on ACEI/ARB have similar mortality and risk of severe illness as compared to patients not taking ACEI/ARB. These results were consistent when analyzed based on type of RAS inhibitors (ACEI or ARB). Among hypertensive patients, ACEI/ARB used appeared to be associated with lower mortality compared to patients not taking ACEI/ARB.

Currently available data from observational studies have shown contrasting findings regarding relationship between ACEI/ARB use and outcomes in Covid-19 patients. In a study of 1128 hospitalized patients with HTN, Zhang et al showed that inpatient use of ACEI/ARB was associated with decreased risk of mortality compared with ACEI/ARB non-users [6]. However, Reynolds et al found no significant association between use of ACEI/ARB and risk of developing critical illness among those tested positive for Covid-19 [7]. Similarly, ACEI/ARB use was not associated with risk of in-hospital mortality in a large multicenter study of hospitalized Covid-19 patients [10]. On the contrary, there was increased percentage of patients admitted to ICU in the ACEI/ARB group compared with non-users in a large multi-center study [11].

In this context, our meta-analysis including >23,000 patients reconciles the findings of existing studies and shows that ACEI/ARB use is not associated with increased risk of mortality or severe illness among a broad patient population with Covid-19. In patients with HTN, our finding of reduced mortality associated with ACEI/ARB use is concordant with few other retrospective studies [6, 18]. We also noted lower proportion of severe illness in ACEI/ARB group compared with non ACEI/ARB group among the HTN patients, although this didn’t reach statistical significance. However, these associations in HTN sub-group are hypothesis generating and should be interpreted cautiously as the analysis was based on studies limited to 1,895 patients. Of relevance, recent studies have suggested a lower SARS-Cov-2 viral load and inflammatory marker levels in hypertensive patients taking ACEI/ARB compared to other anti-hypertensive medications (14, 18). Further, an association between ACEI/ARB use and increased risk of hospital admission or severe illness in previous studies is likely a result of confounding bias due to relative incidence of CV comorbidities.

With lack of randomized controlled trials (RCTs) to date, our findings have important implications for the management of Covid-19 patients with CV disease. SARS-CoV-2 mediated downregulation of ACE2 and immune dysregulation with endothelial dysfunction, myocardial injury and prothrombotic state might lead to a downward spiral thus accounting for worse outcomes in these patients [21]. ACE2 metabolizes angiotensin II into angiotensin 1-7 that serves to counter regulate the deleterious effects of RAS. While the role of ACE2 as cellular receptor for SARS-CoV-2 entry into host cells is proven, human studies have been inconclusive with respect to the effect of RAS inhibitors on ACE2 levels [22]. Furthermore, contradictory to the speculation that RAS inhibitors might increase risk of infection via upregulation of ACE2, the postulated effects on ACE2 expression have been actually suggested to protect against severe lung injury in these patients [23]. In the absence of clinical data, however, human studies are needed to further explore this complex interaction between RAS inhibitors and ACE2, particularly in lung tissues.

There are several limitations to our study. First, our pooled analyses were based on observational studies that have inherent risk of bias due to confounding variables. Patients taking ACEI/ARBs have increased burden of other comorbidities that might make them more prone to fatality. However, despite this potential bias, we observed no association of ACEI/ARB use with increased risk of mortality or severe illness. Second, ascertainment of drug data is limited in individual retrospective studies. It remains unknown whether continuation or withdrawal of these drugs during hospitalization influenced outcomes in patients admitted with Covid-19.

In conclusion, our study provides reassurance that there is no increased risk of mortality or severe illness in patients using ACEI/ARB compared to non-users. Given the high prevalence of CV disease and complications in Covid-19, continuation of RAS inhibitors should be encouraged and eventually driven by clinical judgment. On-going RCTs will further inform the safety and efficacy of these drugs in HTN patients with Covid-19 disease.

## Data Availability

Not applicable

**Abbreviations**
ACE: Angiotensin converting enzyme
ACEI: ACE inhibitors
ARB: Angiotensin receptor blockers
CI: Confidence intervals
CV: Cardiovascular
HTN: Hypertension
OR: Odds ratio
RAS: Renin angiotensin system

## References

1) Zhu N, Zhang D, Wang W, et al; China Novel Coronavirus Investigating and Research Team. A novel coronavirus from patients with pneumonia in China, 2019. N Engl J Med. 2020;382(8):727–733. doi:10.1056/NEJMoa2001017

2) Li W, Moore MJ, Vasilieva N, et al. Angiotensin-converting enzyme 2 is a functional receptor for the SARS coronavirus. Nature 2003; 426: 450–4.

3) Ferrario CM, Jessup J, Chappell MC, et al. Effect of angiotensin-converting enzyme inhibition and angiotensin II receptor blockers on cardiac angiotensin-converting enzyme 2. Circulation 2005; 111: 2605-10.

4) Messerli FH, Siontis G, Rexhaj E. COVID-19 and Renin Angiotensin Blockers: Current Evidence and Recommendations. Circulation. 2020 Apr 13. doi: 10.1161/CIRCULATIONAHA.120.047022.

5) Guo T, Fan Y, Chen M, et al. Cardiovascular implications of fatal outcomes of patients with coronavirus disease 2019 (COVID-19). JAMA Cardiol 2020 March 27 (Epub ahead of print).

6) Zhang P, Zhu L, Cai J, et al. Association of Inpatient Use of Angiotensin Converting Enzyme Inhibitors and Angiotensin II Receptor Blockers with Mortality Among Patients With Hypertension Hospitalized With COVID-19. Circ Res. 2020 Apr 17. doi: 10.1161/CIRCRESAHA.120.317134.

7) Reynolds HR, Adhikari S, Pulgarin C, et al. Renin-Angiotensin-Aldosterone System Inhibitors and Risk of Covid-19. N Engl J Med. 2020 May 1. doi: 10.1056/NEJMoa2008975.

8) Moher D, Shamseer L, Clarke M, et al; PRISMA-P Group. Preferred reporting items for systematic review and meta-analysis protocols (PRISMA-P) 2015 statement. Syst Rev 2015;4:1.

9) Peng YD, Meng K, Guan HQ, et al. Clinical Characteristics and Outcomes of 112 Cardiovascular Disease Patients Infected by 2019-nCoV. Zhonghua Xin Xue Guan Bing Za Zhi. 2020 Mar 2;48(0):E004.

10) Mehra MR, Desai SS, Kuy S, Henry TD, Patel AN. Cardiovascular Disease, Drug Therapy, and Mortality in Covid-19. N Engl J Med. 2020 May 1. doi: 10.1056/NEJMoa2007621.

11) Mehta N, Kalra A, Nowacki AS, et al. Association of Use of Angiotensin-Converting Enzyme Inhibitors and Angiotensin II Receptor Blockers With Testing Positive for Coronavirus Disease 2019 (COVID-19). JAMA Cardiol. 2020 May 5. doi: 10.1001/jamacardio.2020.1855.

12) Mancia G, Rea F, Ludergnani M, Apolone G, Corrao G. Renin-Angiotensin-Aldosterone System Blockers and the Risk of Covid-19. N Engl J Med. 2020 May 1. doi: 10.1056/NEJMoa2006923.

13) Li J, Wang X, Chen J, Zhang H, Deng A. Association of Renin-Angiotensin System Inhibitors With Severity or Risk of Death in Patients With Hypertension Hospitalized for Coronavirus Disease 2019 (COVID-19) Infection in Wuhan, China. JAMA Cardiol. 2020 Apr 23. doi: 10.1001/jamacardio.2020.1624.

14) Yang G, Tan Z, Zhou L, et al. Effects Of ARBs And ACEIs On Virus Infection, Inflammatory Status And Clinical Outcomes In COVID-19 Patients With Hypertension: A Single Center Retrospective Study. Hypertension. 2020 Apr 29. doi: 10.1161/HYPERTENSIONAHA.120.15143.

15) Feng Y, Ling Y, Bai T, et al. COVID-19 with Different Severity: A Multi-center Study of Clinical Features. Am J Respir Crit Care Med. 2020 Apr 10. doi: 10.1164/rccm.202002-0445OC.

16) Wang Y, Lu X, Chen H, et al. Clinical Course and Outcomes of 344 Intensive Care Patients with COVID-19. Am J Respir Crit Care Med. 2020 Apr 8. doi: 10.1164/rccm.202003-0736LE.

17) Li X, Xu S, Yu M, et al. Risk factors for severity and mortality in adult COVID-19 inpatients in Wuhan. J Allergy Clin Immunol. 2020 Apr 12. pii: S0091–6749(20)30495-4.

18) Meng J, Xiao G, Zhang J, et al. Renin-angiotensin system inhibitors improve the clinical outcomes of COVID-19 patients with hypertension. Emerg Microbes Infect. 2020 Dec;9(1):757–760.

19) de Abajo FJ, Rodríguez-Martín S, Lerma V, et al; MED-ACE2-COVID19 study group. Use of renin-angiotensin-aldosterone system inhibitors and risk of COVID-19 requiring admission to hospital: a case-population study. Lancet. 2020 May 14:S0140–6736(20)31030-8. doi: 10.1016/S0140-6736(20)31030-8.

20) Conversano A, Melillo F, Napolano A, et al. RAAs inhibitors and outcome in patients with SARS-CoV-2 pneumonia. A case series study. Hypertension. 2020 May 8. doi: 10.1161/HYPERTENSIONAHA.120.15312. Epub ahead of print.

21) Madjid M, Safavi-Naeini P, Solomon SD, Vardeny O. Potential Effects of Coronaviruses on the Cardiovascular System: A Review. doi:10.1001/jamacardio.2020.1286

22) Vaduganathan M, Vardeny O, Michel T, McMurray J, Pfeffer MA, Solomon SD. Renin–Angiotensin–Aldosterone System Inhibitors in Patients with Covid-19. N Engl J Med 2020; 382: 1653–1659.

23) Kuba K, Imai Y, Rao S, et al. A crucial role of angiotensin converting enzyme 2 (ACE2) in SARS coronavirus-induced lung injury. Nat Med. 2005;11 (8):875–879. doi:10.1038/nm1267.

